# Micronutrient Knowledge, Attitudes, and Dietary Practices Among Women of Reproductive Age in Aden, Yemen: A Cross-Sectional Study

**DOI:** 10.64898/2026.10.05.26364809

**Authors:** Mohammed A. A. Al-Baghdadi, Shifaa Suhail Mohammed, Shrook Jalal Ahmed, Mohammed Ali Khalifa

**Affiliations:** Assoc. Prof. MD in Clinical Microbiology& Immunology, Department of Human Medicine, Faculty of Medicine and Health Sciences, University of Science and Technology, Aden, Yemen; Assoc. Prof. MD in Clinical Microbiology & Immunology, Department of Paraclinic, Faculty of Medicine & Health Sciences, Aden University; Bachelor of Clinical Nutrition and Dietetic, Department of Health sciences, Faculty of Medicine and Health Sciences, University of Science and Technology, Aden, Yemen; Pharmacy Department, Faculty of Medicine and Health Sciences, University of Science and Technology, Aden, Yemen; Faculty of Pharmacy, Omdurman Islamic University, Khartoum, Sudan

**Keywords:** Micronutrients, Knowledge, Attitudes, Practices, Women of Reproductive Age, Yemen, Dietary diversity

## Abstract

**Background:** Micronutrient deficiencies are a major public health problem for women of reproductive age (WRA), especially in conflict settings, such as Yemen. However, there is little information on knowledge, attitudes and practices (KAP) toward micronutrient-rich foods among WRA in Aden Governorate. This study aimed to assess knowledge, attitude and practices of WRA toward micronutrient rich foods attending health facilities in Aden Governorate, Yemen.

**Methods:** A facility based descriptive cross-sectional study was conducted among 348 WRA aged 15-49 years who visited selected primary health care and maternal and child health facilities in the eight districts of Aden governorate, Yemen. Data were collected using structured interviewer administered questionnaire which was adapted from guidelines of Food and Agriculture Organization and World Health Organization. Chi-square test and Pearson correlation were used to assess the association between the KAP indicators and sociodemographic characteristics.

**Results:** The mean age of the participants was 26.3 ± 8.8 years and 73.9% had high-grade education. In general, 77.9% identified micronutrients as essential for health and 92.8% correctly identified liver/red meat as micronutrient-rich food. However, only 29.6% identified iodized salt as a source of iodine. Although 95.7% perceived supplementation as important, only 9.8% reported regular supplement use. The most commonly reported barrier to access to nutritious food was high food cost (43.4%) and 70.6% perceived healthy foods as too expensive. Restriction in daily consumption was observed for eggs (36.2%), meat/poultry/fish (32.2%) and fruits (13.2%). Education was moderately positively correlated with micronutrient knowledge (r = 0.514, p < 0.001) and age was weakly positively correlated (r = 0.162, p = 0.002).

**Conclusion:** WRA in Aden had good knowledge and positive perceptions about micronutrients and supplementation but major gaps in dietary practices and access to affordable nutritious foods. Integrated, culturally appropriate interventions that include nutrition education and improved access to affordable micronutrient-rich foods and appropriate supplementation are needed.

## 1. Introduction

Food insecurity is a major global driver of poor health and nutritional vulnerability and has serious consequences for the health of individuals and populations. The effects are seen in different forms of malnutrition, such as undernutrition, overnutrition, and deficiencies in essential macronutrients and micronutrients, especially where there is little dietary variety (1,2,1). In this context, the nutritional status of women is an important indicator of general well-being of society (3) and is closely associated with the nutritional security and health of their children (4). Adequate nutrition during the reproductive years is therefore crucial to preserving maternal health, reproductive function and optimizing pregnancy and birth outcomes. However, insufficient intake of food at this crucial stage of life can lead to a lack of vital nutrients. The World Health Organization has observed that suboptimal intake of micronutrients in women of reproductive age can have intergenerational consequences, affecting the health and well-being of future generations (5).

Yemen is one of the most complex humanitarian contexts in which these nutritional vulnerabilities are experienced. “Prolonged conflict, economic decline, displacement and disruption of food and health systems have led to a dire food and nutrition crisis affecting over 20 million people, including around 10 million facing acute food insecurity. Women and children are especially vulnerable to the nutritional impacts of this crisis, bearing a disproportionate share of malnutrition. Serious consequences such as impaired physical and cognitive development in children and increased risk of morbidity and mortality may result from such nutritional deprivation (6,7). Moreover, the huge displacement, socioeconomic instability and ongoing violence have resulted in a high burden of gender-based violence on women and girls, further compounding their social and health vulnerabilities (8).

Micronutrient deficiencies represent a substantial part of the nutritional burden in women in Yemen and other low- and middle-income settings. In Yemen, 38% of pregnant women have iron-deficiency anemia, indicating the level of iron-related nutritional problems among women of reproductive age (9). Deficiencies in iron, vitamin A and folic acid are of particular concern due to their possible adverse effects on maternal health, pregnancy outcomes and child health (10). The problem of micronutrient deficiency is exacerbated by poor nutrition-related knowledge. Evidence from Yemen shows that only 7.1% of women have adequate knowledge of iron-deficiency anemia, indicating a significant knowledge gap that may impede the adoption of appropriate dietary and preventive practices (9).

Further evidence from Aden and other parts of Yemen supports the poor dietary diversity of women of reproductive age. In 2024, a study done in the Sheik Othman district of Aden reported low prevalence of minimum dietary diversity among women of reproductive age. Similarly, evidence from Sana’a governorate showed that more than 71% of women of reproductive age consumed food from less than five food groups (11,12). These results reflect a lack of dietary variety and suggest limited access to a variety of nutrient-rich foods. The ongoing humanitarian crisis is a significant contextual determinant, as economic hardship, food insecurity and disruption of food systems can severely limit household access to nutritious foods. Thus, poor dietary diversity could increase women’s susceptibility to micronutrient deficiency and its associated poor health outcomes such as anemia (13).

In an effort to achieve optimal nutritional status, reproductive health and well-being, for women of reproductive age, a balanced and diverse diet is vital. Food items such as fish, meat, legumes, leafy green vegetables, citrus fruits and nuts may be significant sources of vital nutrients in the traditional dietary patterns of Aden. However, economic, social and cultural factors may exert a considerable influence on the ability of women and households to adopt nutritionally adequate dietary patterns (14,11). The availability of potentially nutrient-rich traditional foods therefore does not necessarily translate into adequate consumption, especially in a context of poverty, food insecurity and limited access to a diversity of foods.

Furthermore, international evidence indicates that nutritional knowledge and attitudes do not automatically translate into appropriate dietary practices. Women of reproductive age in marginalized groups in Sri Lanka have shown poor nutritional knowledge despite the generally positive attitudes toward receiving nutrition information (15). Similarly, studies among female students in Malaysia found that knowledge and attitudes regarding folate intake were relatively good; however, this did not necessarily translate into proper dietary practices which may be attributed to their life stage and lack of immediate pregnancy-related concerns (16). In Sudan, women have been found to have moderate intake of essential micronutrients, and children have been found to have substantially inadequate micronutrient consumption, pointing to continuing nutritional vulnerabilities across population groups (17). Together, these findings suggest that improving micronutrient intake cannot be simply a matter of making nutritional recommendations available, but must account for the complex interaction of knowledge, attitudes, practices, socioeconomic circumstances and the wider food environment.

Furthermore, the persistent high prevalence of micronutrient deficiencies, such as anemia, in Asian populations calls into question the effectiveness and delivery of existing strategies for food fortification and micronutrient supplementation (18). Supplementation and fortification continue to be important public health approaches but their effectiveness may be limited if underlying determinants are not effectively addressed. These determinants include, but are not limited to, inadequate dietary diversity, poor nutrition knowledge, socioeconomic constraints, cultural practices and barriers to access to health and nutrition services.

In this context, assessment of knowledge, attitudes and practices (KAP) of women of reproductive age on micronutrient-rich foods is particularly important. Understanding these dimensions can help to identify modifiable behavioral and informational factors contributing to inadequate micronutrient intake and provide an evidence base for designing context-specific interventions. Although there are evidence of the burden of food insecurity and poor dietary diversity and micronutrient deficiencies in Yemen, limited data are available on KAP regarding micronutrient-rich foods among women of reproductive age in Aden. Thus, the present study addresses a significant gap in evidence by investigating these determinants in a conflict-affected urban context. The findings could provide a basis for developing culturally appropriate nutrition education programs and evidence-based public health strategies to improve dietary practices and reduce micronutrient deficiencies, especially iron and iodine deficiencies, among women of reproductive age.

Serious public health concerns related to micronutrient deficiencies among women of reproductive age in Aden, Yemen are being driven by a variety of interrelated factors associated with the ongoing humanitarian crisis. Food insecurity, disruption of food systems, limited access to nutrient-dense foods, socioeconomic constraints, cultural beliefs and food taboos, poor nutrition awareness and weaknesses within the health system may all contribute to inadequate dietary diversity and micronutrient intake. These factors may have negative implications on women’s nutritional and reproductive health and may also perpetuate an intergenerational cycle of malnutrition; whereby maternal nutritional deficiencies increase the vulnerability of future generations.

This study aims to address the significant global burden of micronutrient deficiencies, especially in women of reproductive age. Micronutrient deficiencies, often called “hidden hunger”, may negatively impact women’s health and reproductive outcomes and may be involved in anemia, and adverse fetal and neonatal outcomes. In contexts like Aden governorate, food insecurity, poverty, existing dietary patterns, cultural practices and limited nutrition awareness may combine to reduce dietary diversity and increase vulnerability to micronutrient deficiencies. Furthermore, lack of knowledge and perhaps negative attitudes toward the appropriate nutrition may be other barriers to adequate consumption of micronutrient-rich foods. Therefore, generating local evidence on these factors is critical to inform targeted nutrition education, preventive interventions and contextually appropriate public health policies.

The aim of this study was to assess the knowledge, attitude and practices (KAP) of micronutrient rich foods among women of reproductive age (WRA) attending primary health centers (PHCs) and maternal and child health (MCH) centers in Aden governorate, Yemen with special focus on nutrition-related behaviors relevant to the promotion of nutritional well-being and prevention of diet-related health problems.

## 2. Materials and Method

### 2.1. Design and Setting of Study

A facility based descriptive cross-sectional study was conducted among women of reproductive age (WRA) of 15-49 years of age attending selected health facilities in Aden Governorate, Yemen. The study was conducted in the eight administrative districts of Aden Governorate including Primary Health Care (PHC) centers, Maternal and Child Health (MCH) centers and Al-Sadaqa General Teaching Hospital. Eligible women were recruited when they presented these facilities for consultation or other health care services during the study period.

### 2.2. Study Population and Inclusion Criteria

The study population was composed of WRA aged 15–49 years who attended the selected health facilities during the study period. Women residing in Aden Governorate and women from surrounding rural areas who had lived in the study area for a period of at least six months were eligible to participate. Women were eligible if aged 15–49 years, lived in the study area for at least six months, visited one of the selected health facilities during the data-collection period, and provided informed consent to participate. Women who refused to participate were excluded. Women who were ill and pregnant, and women who were acutely ill or seeking care for an illness at the time data were collected were also excluded.

### 2.3. Sample size

The sample size required was calculated using the standard formula for estimating a single population proportion:

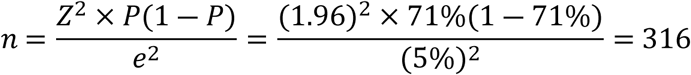

Where:

- *Z* = 1.96 for 95% confidence.
- *p* = The Prevalence is based on a study conducted in Sheikh Othman district of Aden (2024), which indicates a low prevalence of minimum dietary diversity among women of reproductive age, and another study conducted in Sana’a governorate showed that 71% of women of reproductive age (WRA) consumed fewer than five food groups (11,12).
- *e* = margin of error accepted (e.g., 0.05).

The sample size was estimated to be 316 participants based on the chosen prevalence estimate. To account for possible non-response and incomplete data, a further 10% was added to give a final target sample size of 348 women.

### 2.4. Sampling Strategy

The study participants were selected using a multistage stratified random sampling technique. In the first stage, health facilities were randomly selected from the eight districts of Aden governorate. In the second stage, the estimated sample size was distributed to the selected health facilities proportionally based on the usual client attendance or caseload. At the last stage, eligible WRA who had visited the selected facilities were recruited using systematic random sampling until the estimated sample size was reached. This was done to ensure representation of women attending different types of health facilities and to minimize potential selection bias associated with recruiting participants from a single facility or district.

### 2.5. Data Collection Tool

Data were collected using a structured questionnaire administered by an interviewer and adapted from previously validated Knowledge, Attitude and Practice (KAP) tools and relevant international guidance developed by the Food and Agriculture Organization (FAO) (19) and the World Health Organization (WHO) (20). Then the questionnaire was modified according to the dietary pattern, available foods and sociocultural context of Aden governorate, Yemen.

The questionnaire was initially developed in English and then translated into Arabic for use with study participants. The questionnaire was reviewed and pilot tested by two nutrition specialists; Professor Somiya Gutbi Salim Mohamed and Associate Professor Dr. Manal Abdeen Mohamed Ali for content, clarity, cultural appropriateness and feasibility. The questionnaire was also pilot tested with 30 WRA who were not included in the final study sample. The feedback from the pilot testing was used to refine the wording, the sequence and clarity of the questionnaire items before the commencement of the main data collection.

### 2.6. Study Variables and KAP Evaluation

The questionnaire included sociodemographic characteristics, household food security, knowledge about micronutrients and their dietary sources, awareness of the consequences of micronutrient deficiencies, attitudes toward micronutrient-rich foods, and dietary practices.

The knowledge section assessed participants’ knowledge of micronutrients, their dietary sources and health consequences related to micronutrient deficiencies. The attitude section looked at perceptions and attitudes toward intake of micronutrient rich foods including the impact of food cost, family preferences and social factors. The practice section evaluated dietary behaviors, including meal practices, consumption of different food groups during the week, use of iodized salt and micronutrient supplementation.

Composite knowledge, attitude and practice scores were obtained by summing responses to the relevant items. The computed scores were utilized to classify the participants based on their KAP levels.

### 2.7. Ethical Considerations

Participants were recruited from 01/06/2026 to 01/09/2026. Before participating all participants gave verbal informed consent. The participants were informed about the aim and procedures of the study and participation was voluntary. Confidentiality and privacy were maintained during the study.

Ethical approval for the study was obtained from the University Research Committee with approval number (UST-ETH-2026-0015). Written informed consent was obtained from all adult participants before participation. For participants under 18 years of age, written informed consent was obtained from their parent or legally authorized guardian before their participation in the study. Participation was voluntary. The participants were informed about the purpose and procedures of the study, and during the process of research, confidentiality and privacy were maintained. The data were collected and used only for research purposes.

### 2.8. Statistical Analysis

The data were entered, coded, and analyzed using the Statistical Package for Social Sciences (SPSS) version 22. Characteristics of the study population were summarized using descriptive statistics. Categorical variables were summarized as frequencies and percentages. Continuous variables were summarized as means and appropriate measures of dispersion.

The scores of knowledges, attitude and practice were calculated according to the scoring criteria predetermined for each domain of the questionnaire. The chi-square (χ2) test was used for the assessment of the associations between categorical variables and KAP categories. Statistical significance was defined as p ≤ 0.05.

## 3. The Results

### 3.1. Participants’ Characteristics

The study included a total of 348 women of reproductive age (WRA). The mean age of participants was (26.3 ± 8.8) years; the age range was (16–47) years. The majority of participants were single (60.1%), had a high education level (73.9%) and were students (53.2%). Almost half of the sample (49.7%) lived in households with 5–7 persons, while 46.0% of them had 1–2 children below five years old. The most frequently reported household sources of income were daily labor (37.9%) and salaried employment (37.6%). Students were the largest occupational group (53.2%) followed by housewives (24.1%) among the study participants.

### 3.2. Awareness of micronutrients and micronutrient-rich foods

Participants showed a relatively high level of recognition of micronutrient-rich foods, but knowledge of individual food sources varied substantially (Table 1). In general, 77.9% of participants agreed that micronutrients are important for health.

**Table 1:** Knowledge of Micronutrients (n=348).

| Indicator | Percentage |
| --- | --- |
| Knew micronutrients are essential for health | 77.9% |
| Correctly identified **liver/red meat** as rich | 92.8% |
| Correctly identified **eggs** as rich | 91.4% |
| Correctly identified **fruits** as rich | 90.5% |
| Correctly identified **dairy** as rich | 89.4% |
| Correctly identified **dark leafy vegetables** as rich | 88.5% |
| Correctly identified **fish/seafood** as rich | 79.3% |
| Correctly identified **legumes** as rich | 77.0% |
| Correctly identified **nuts/seeds** as rich | 75.9% |
| Correctly identified **iodized salt** as rich | **29.6%** |

Animal-source foods were generally acknowledged as key sources of micronutrients. (92.8%) of participants correctly identified liver and red meat, followed by eggs (91.4%), fruits (90.5%), dairy products (89.4%), and dark green leafy vegetables (88.5%). Fish and seafood were recognized by 79.3%, while legumes and nuts/seeds were identified by 77.0% and 75.9%, respectively. Whereas iodized salt was correctly identified as a relevant source of micronutrients by 29.6%.

Knowledge of the effects of micronutrient deficiencies was much less comprehensive. Only 26.2% of the participants were able to identify all major deficiency-related consequences assessed in the questionnaire, such as anemia, osteoporosis, night blindness, fatigue, weakened immunity and goiter. Micronutrient deficiencies and their consequences were unknown to 6.9% and 9.5%, respectively, while 9.5% and 6.9%, respectively, identified anemia alone. Concerning the importance of micronutrient-rich foods, 46.0% of the participants perceived these foods to be predominantly important for the health of infants, whereas 31.6% recognized their importance for both maternal and infant health. A further 18.1% felt that they were mostly beneficial for the women’s own health. 4.3% said they did not know, as shown in Table 2.

**Table 2.** Perceived Importance of Micronutrient-Dense Foods for Maternal and Child Health Among Women of Reproductive Age (n = 348).

| **The Importance of WRA to Eat Foods Rich in Micronutrients** | **n(%)** |
| --- | --- |
| Excellent for her infant | 160(46%) |
| Excellent for both Health | 110(31.6%) |
| Excellent for her Health | 63(18.1%) |
| Don't Know | 15(4.3%) |

### 3.3. Overall KAP Scores

The total KAP assessment showed significant differences in the three domains (Table 3). The mean knowledge score was 87.3%, indicating a high level of knowledge according to the scoring system of the study. Specifically, 95.2% of participants agreed that dietary diversity is important and 96.5% recognized that pregnant women need more nutritious food, as presented in Fig. 1.

**Fig. 1:**
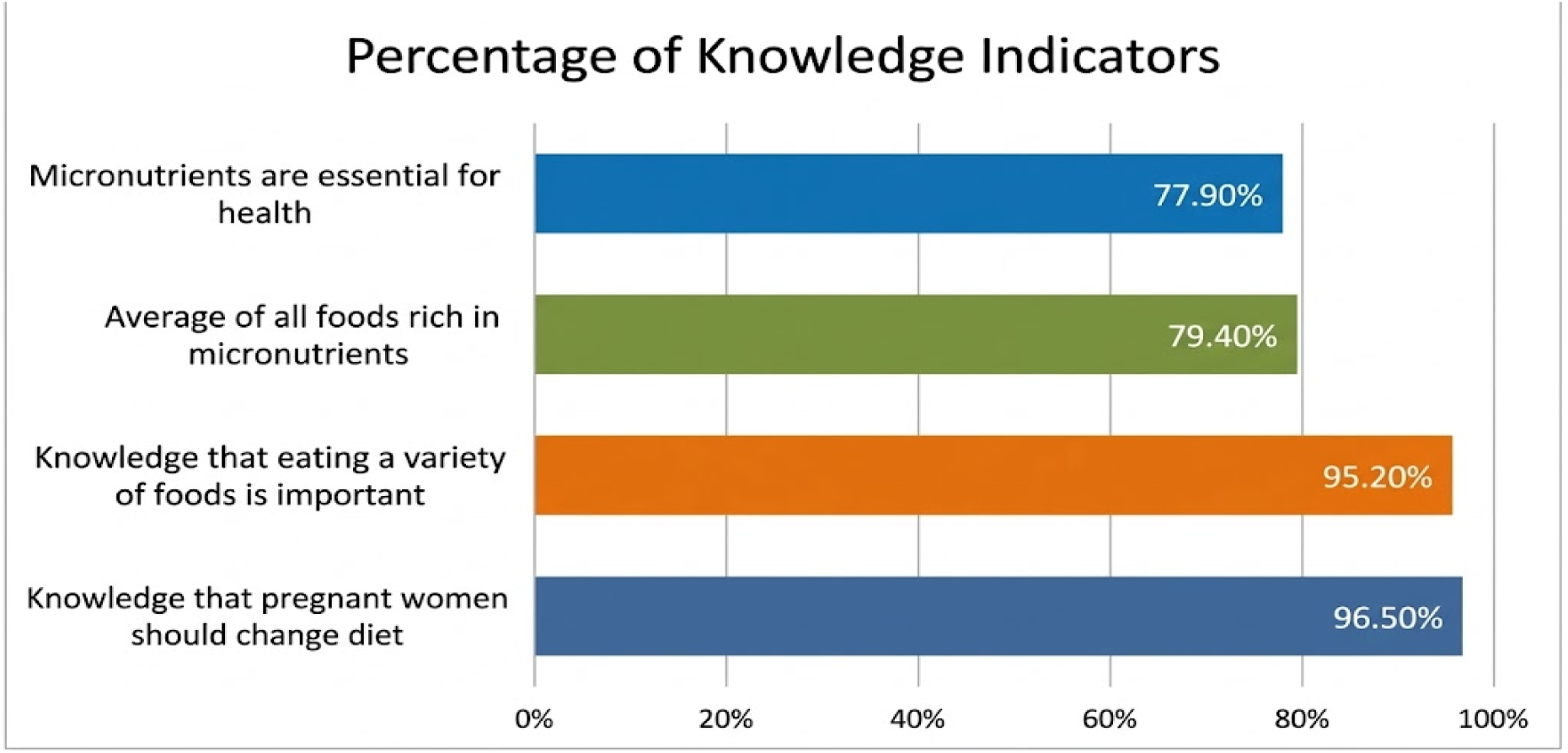
The average percentage of knowledge indicators

**Table 3:** Knowledge, Attitudes, and Practices (KAP) Related to Nutritious Food Consumption Among Women of Reproductive Age (n=348).

| **Domain** | **Score** | **Main Findings** |
| --- | --- | --- |
| **Knowledge** | **87.3%** | 95.2% agreed dietary diversity was important; 96.5% knew pregnant women need more nutritious foods. |
| **Attitude** | **19.5%** | 70.6% perceived healthy food as too expensive; 59.5% preferred traditional foods; 32.7% reported social influence on cooking. |
| **Practice** | **71.6%** | 67.2% confident preparing healthy meals on a budget; weekly consumption of food groups varied. |

Overall attitude score was 19.5% compared to 17.5%. A majority of the participants (70.6%) felt that healthy food was expensive. 59.5% of the participants preferred traditional foods to healthier options. Additionally, 32.7% stated social influences affected food preparation and dietary choices, as shown in Fig. 2.

**Fig. 2:**
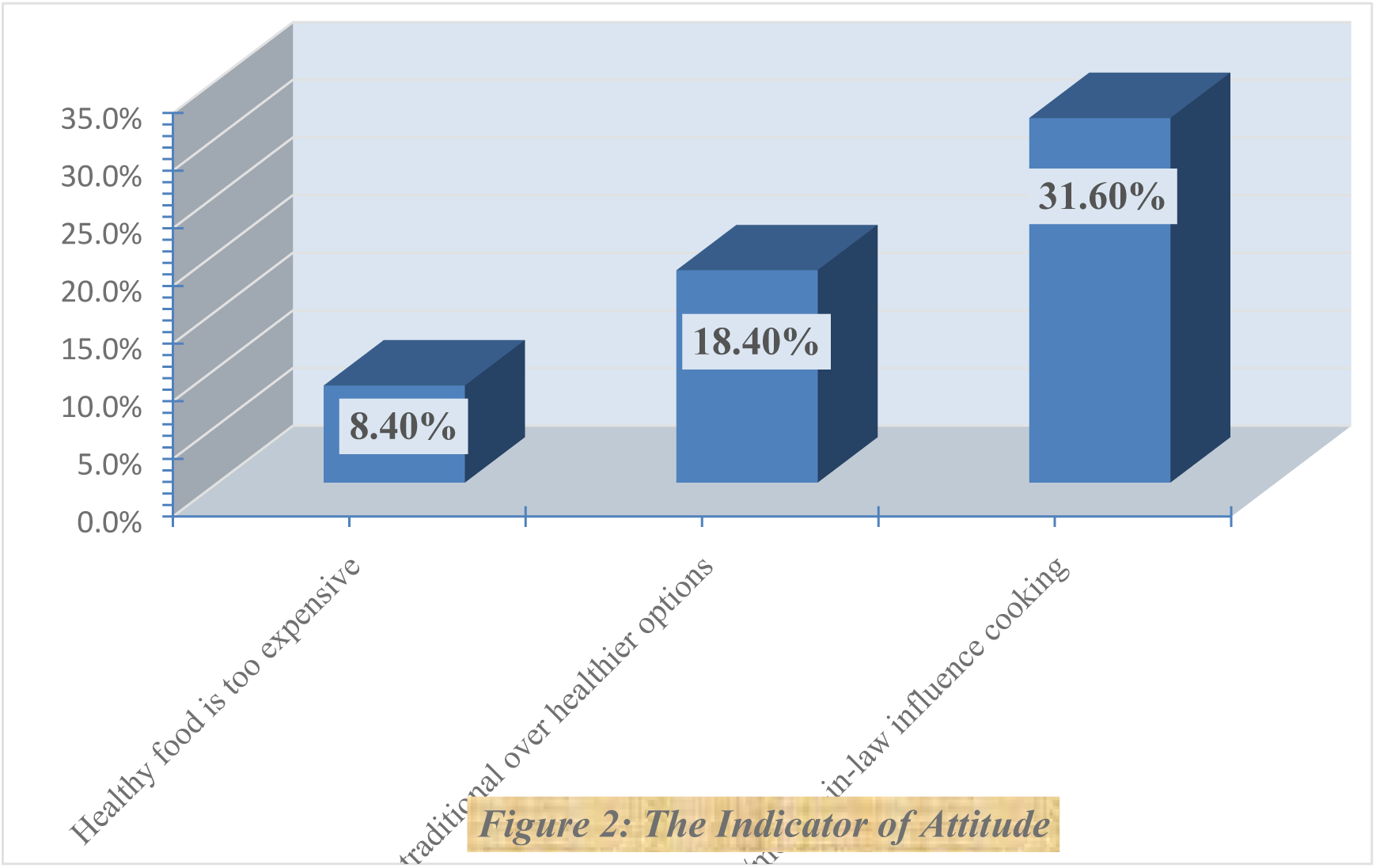
The average percentage of attitude indicators

The overall practice score was 71.6%. Roughly 67.2% of the participants felt confident in preparing healthy meals on a limited budget. Other practice indicators were use of iodized salt, intake of different food groups and micronutrient supplementation, as described in Fig. 3.

**Fig. 3:**
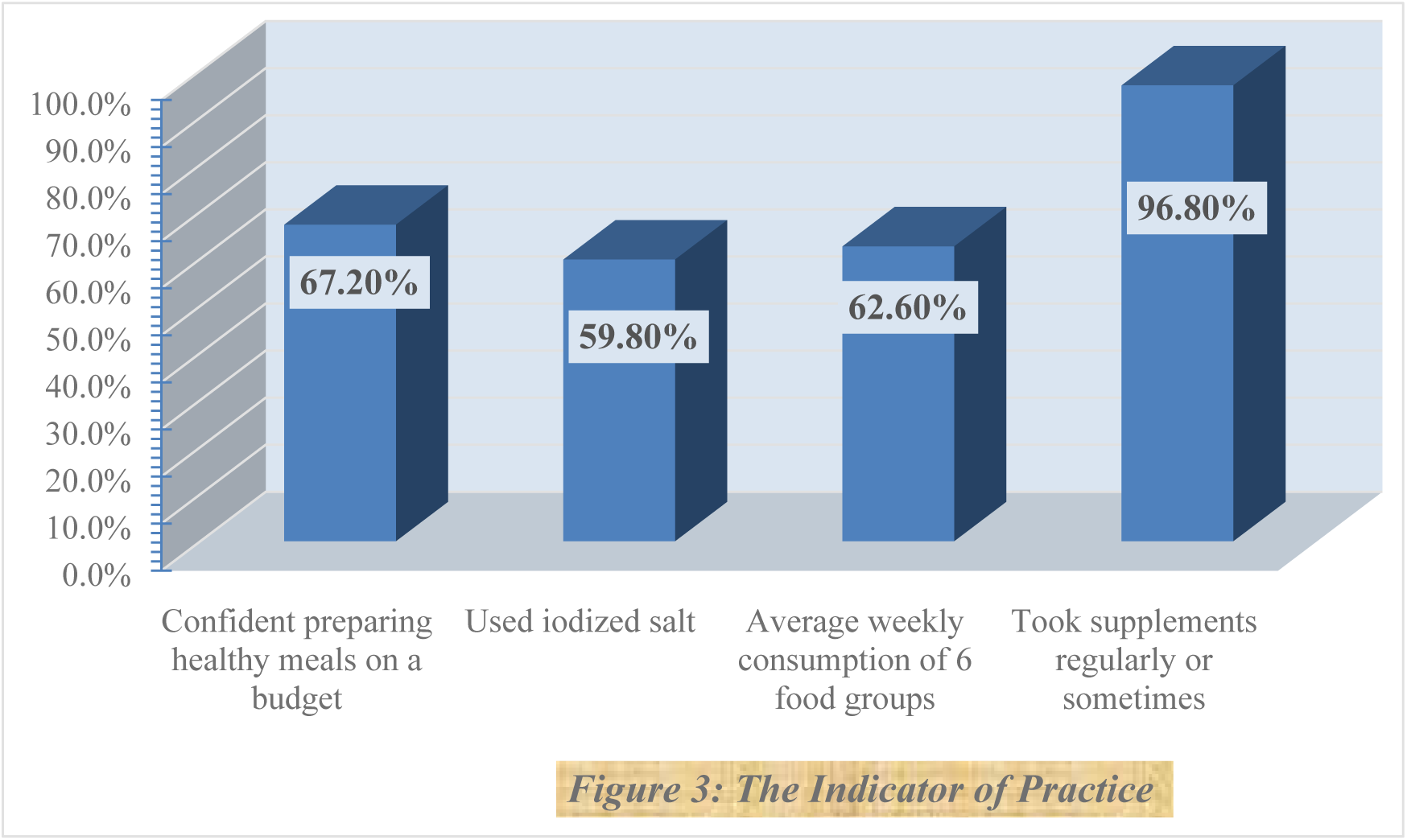
The average percentage of practice indicators

### 3.4. Eating Patterns

Majority of the participants reported eating three meals per day (76.2%) while 22.4% reported eating two meals per day and 1.4% reported eating more than three meals daily (Table 4). Most frequently, breakfast was consumed at 7:00 AM (34.2%). Roti with egg (30.2%) was the most frequently reported breakfast food, and tea was the most commonly reported breakfast beverage (49.1%). Lunch was most often consumed after 3:00 PM and before 5:00 PM (26.4%). The most commonly reported lunch combination was rice and sanuna 41.1%). The most common meal combination for dinner was roti with Yemeni lahsa/shakshouka (25.0%).

**Table 4:** The Dietary Pattern of Women of Reproductive Age ( n = 3 4 8 ).

| **Meal Pattern** | **N (%)** |
| --- | --- |
| 3 meals/day | 265 (76.2%) |
| 2 meals/day | 78 (22.4%) |
| > 3 Meals | 5 (1.4%) |
| **Breakfast** | Most common: 7:00 AM (34.2%); food: Roti & Egg (30.2%); drink: Tea (49.1%) |
| **Lunch** | Peak: after 3–5 PM (26.4%); food: Rice & Sanuna (41.1%) |
| **Dinner** | Most common: Roti with Yemeni Lahsa/Shakshouka (25.0%) |

### 3.5. Weekly Consumption of Food Groups

There was a lot of variation in how often different food groups were eaten (Table 5). Eggs were the most commonly consumed food group, with 75.8% of participants consuming them on at least four days a week and 36.2% consuming them daily. 71.8% of participants consumed legumes ≥ 4 days/week and 31.9% of participants consumed them daily. Meat, poultry and fish were eaten at least four days per week by 61.8% of participants and every day by 32.2%. 61.5% of the participants consumed dark green leafy vegetables at least four days per week and 27.6% reported daily consumption. 57.8% reported eating dairy products at least 4 days per week and 21.0% reported eating dairy products every day. The least consumed food group among those assessed was fruits. Only 46.6% of the participants consumed fruits at least for four days per week, and only 13.2% consumed fruits daily.

**Table 5:** The Weekly Food Group Consumption.

| **Food Group** | **Consumed $\geq 4$ days/week** | **Consumed daily** |
| --- | --- | --- |
| Eggs | 75.8% | 36.2% |
| Legumes | 71.8% | 31.9% |
| Meat/Poultry/Fish | 61.8% | 32.2% |
| Dark leafy vegetables | 61.5% | 27.6% |
| Dairy | 57.8% | 21.0% |
| Fruits | 46.6% | 13.2% |

### 3.6. Salt Iodization and Micronutrient Supplementation

As seen in Table 6, 59.8% of the participants reported use of iodized salt in their households while 21.3% reported use of non-iodized salt. Another 18.9% could not identify the type of salt used. Only 9.8% reported taking supplements regularly in relation to micronutrient supplementation. The majority (87.0%) reported occasional supplement use, 3.2% reported using no supplements.

**Table 6:**
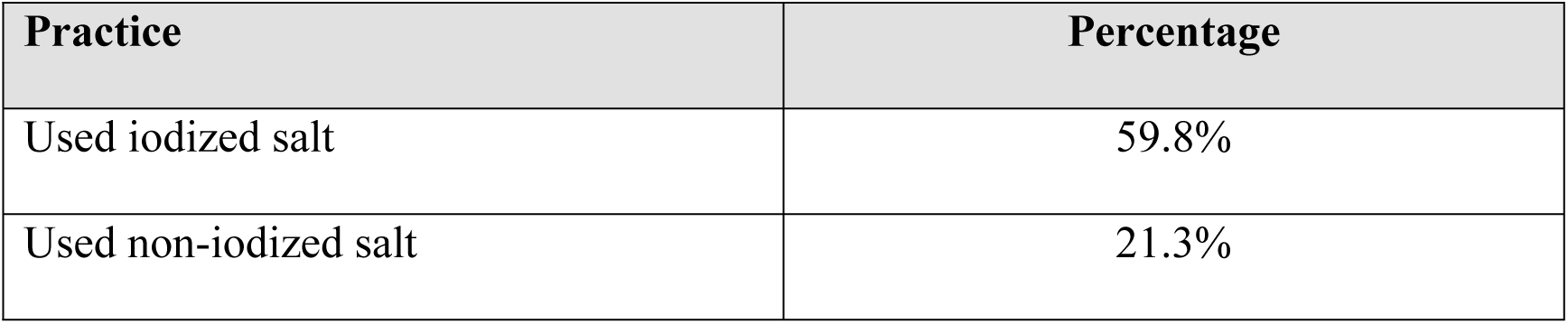
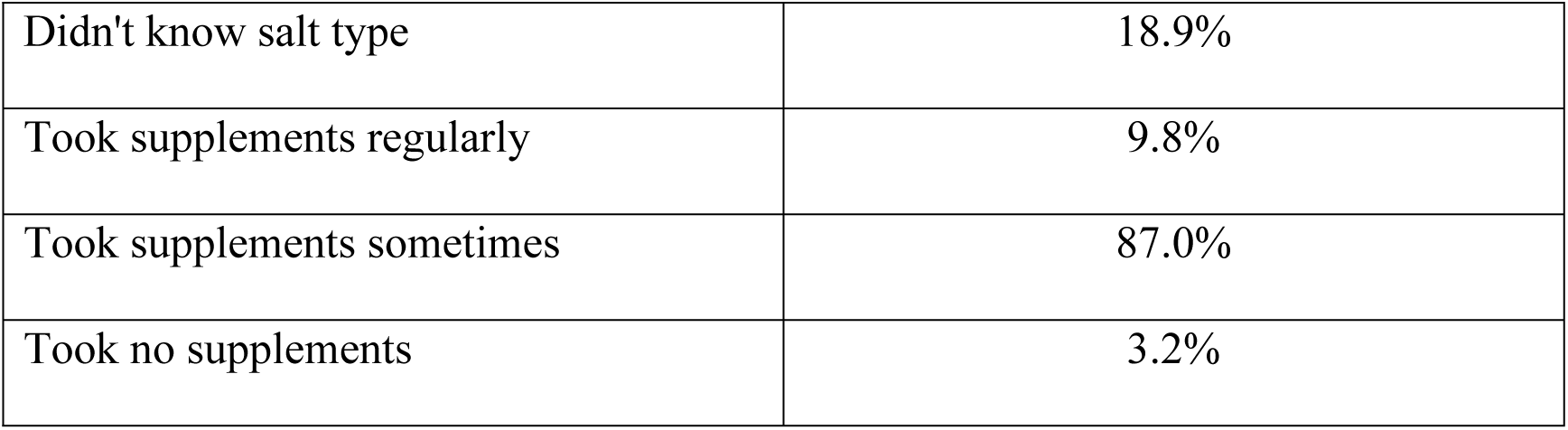
Salt Usage and Supplementation.

| **Practice** | **Percentage** |
| --- | --- |
| Used iodized salt | 59.8% |
| Used non-iodized salt | 21.3% |
| Didn't know salt type | 18.9% |
| Took supplements regularly | 9.8% |
| Took supplements sometimes | 87.0% |
| Took no supplements | 3.2% |

### 3.7. Barriers to Accessing Micronutrient-Dense Foods

The most common barrier to access to nutritious foods was high food cost (43.4%) (Table 7). Other food preferences were reported by 19.8% of the participants. (3.7%) reported not knowing how to prepare food as a barrier.

**Table 7:** The Barriers to Nutritious Food Access of Women of Reproductive Age (n=348).

| Barrier | Percentage |
| --- | --- |
| High cost of food | 43.4% |
| Family preferences for other foods | 19.8% |
| No barriers reported | 8.9% |
| Lack of knowledge on preparation | 3.7% |
| Multiple barriers combined | 3.5% |
| 2 – 3 barriers combined | 20.7% |

Only 8.9% of respondents reported no barriers to access to nutritious foods. 3.5% reported multiple barriers, 20.7% reported two or three barriers simultaneously.

### 3.8. Association of sociodemographic characteristics with knowledge

The relationship between selected sociodemographic characteristics and knowledge of micronutrients was analyzed by chi-square test and Pearson correlation (Table 8). Knowledge differed significantly between age groups (χ², p < 0.001). The highest percentage of participants aware of micronutrients was among those aged 16–20 years (87.1%), followed by those aged 21–25 years (80.5%), >30 years (76.9%) and 26–30 years (51.0%). Pearson correlation indicated a statistically significant positive correlation between age and knowledge score (r = 0.162, p = 0.002).

**Table 8:** The Association Between Sociodemographic Characteristics with the Knowledge about Micronutrients (n=348).

| Grouped Age Years (n) | Knowledge about Micronutrients |  | p-value | r (P-value) |
| --- | --- | --- | --- | --- |
|  | No n(%) | Yes n(%) |  |  |

| 16 – 20 (132) | 17(12.9%) | 115(87.1%) | **0.000** | **0.162(0.002)** |
| --- | --- | --- | --- | --- |
| 21 – 25 (87) | 17(19.5%) | 70(80.5%) |  |  |
| 26 – 30 (51) | 25(49.0%) | 26(51.0%) |  |  |
| > 30 (78) | 18(23.1%) | 60(76.9%) |  |  |
| **Total** | **77(22.1%)** | **271(77.9%)** |  |  |
| **Marital status (n)** | **Knowledge about Micronutrients** |  | **p-value** | **r (P-value)** |
|  | **No n(%)** | **Yes n(%)** |  |  |
| Single (209) | 26(12.4%) | 183(87.6%) | **0.000** | **-0.116-(0.03)** |
| Married (80) | 46(57.5%) | 34(42.5%) |  |  |
| Divorced (49) | 1(2.0%) | 48(98.0%) |  |  |
| Widowed (10) | 4(40.0%) | 6(60.0%) |  |  |
| **Education Level (n)** | **Knowledge about Micronutrients** |  | **p-value** | **r (P-value)** |
|  | **No n(%)** | **Yes n(%)** |  |  |
| Illiterate (13) | 13(100.0%) | 0(0.0%) | **0.000** | **0.514(0.000)** |
| Primary (6) | 5(83.3%) | 1(16.7%) |  |  |
| Secondary (72) | 30(41.7%) | 42(58.3%) |  |  |
| High Grade (257) | 29(11.3%) | 228(88.7%) |  |  |
| **Current Occupation (n)** | **Knowledge about Micronutrients** |  | **p-value** | **r (P-value)** |
|  | **No n(%)** | **Yes n(%)** |  |  |
| Housewife (84) | 45(53.6%) | 39(46.4%) | **0.000** | **0.378(0.000)** |
| Government Employed (30) | 4(13.3%) | 26(86.7%) |  |  |
| Private Employed (45) | 9(20.0%) | 36(80.0%) |  |  |
| Student (185) | 17(9.2%) | 168(90.8%) |  |  |
| Unemployed (4) | 2(50.0%) | 2(50.0%) |  |  |

Knowledge was also significantly different by marital status (p < 0.001). The proportion demonstrating knowledge was 87.6% among single women, as compared with 42.5% among married women, 98.0% among divorced women, and 60.0% among widowed women. The Pearson correlation was statistically significant (r = −0.116, p = 0.030).

Level of education was significantly associated with knowledge on micronutrients (p < 0.001). Based on the dichotomous classification used in this study, 0% of the illiterate’s showed knowledge while 16.7% of the primary education, 58.3% of the secondary education and 88.7% of the high-grade education participants showed knowledge. There was a significant positive association between education level and knowledge as determined by Pearson correlation (r = 0.514, p < 0.001).

There was also a significant difference in knowledge by occupation (p < 0.001). The greatest proportion of knowledge was observed among students (90.8%), government-employed (86.7%) and women employed privately (80.0%). The proportion of participants with knowledge among housewives was 46.4%, and the proportion of participants with knowledge among unemployed participants was 50.0%. Occupation was significantly positively correlated with knowledge (r = 0.378, p < 0.001).

### 3.9. Sociodemographic Characteristics and Perceived Cost of Supplements

We have studied the association between sociodemographic characteristics and the perception that regular supplementation was too expensive using the chi-square test and Pearson correlation in Table 9. There was a statistically significant association between age group and this perception (p = 0.005). The highest percentage of women who reported that supplements were too expensive was in the age group of 21-25 years (14.9%), followed by those in the age group of 26-30 years (13.7%) and 16-20 years (6.1%). This perception was reported by only 1.3% of women aged >30 y. The perception that supplements were too expensive was not statistically significantly associated with marital status (p = 0.091), education level (p = 0.600), or occupation (p = 0.100).

**Table 9:** The Association between Sociodemographic Characteristics and Attitude of WRA about supplements being too expensive to take regularly (n=348).

| Grouped Age Years (n) | Too Expensive to Take Supplements |  | p-value | r (p-value) |
| --- | --- | --- | --- | --- |
|  | No n (%) | Yes n (%) |  |  |
| 16 – 20 (132) | 124(93.9%) | 8(6.1%) | **0.005** | **-0.047-**<br>**(0.4)** |
| 21 – 25 (87) | 74(85.1%) | 13(14.9%) |  |  |
| 26 – 30 (51) | 44(86.3%) | 7(13.7%) |  |  |
| > 30 (78) | 77(98.7%) | 1(1.3%) |  |  |
| **Total (348)** | 319(91.7%) | 29(8.3%) |  |  |
| Marital Status (n) | Too Expensive to Take Supplements |  | p-value | r (p-value) |
|  | No n (%) | Yes n (%) |  |  |
| Single (209) | 188(90.0%) | 21(10.0%) | **0.091** | **-0.116-**<br>**(0.030)** |
| Married (80) | 72(90.0%) | 8(10.0%) |  |  |

| Divorced (49) | 49(100.0%) | 0(0.0%) |  |  |
| --- | --- | --- | --- | --- |
| Widowed (10) | 10(100.0%) | 0(0.0%) |  |  |
| Education Level (n) | Too Expensive to Take Supplements |  | p-value | r (p-value) |
|  | No n (%) | Yes n (%) |  |  |
| Illiterate (13) | 13(100.0%) | 0(0.0%) | **0.6** | **0.034 (0.5)** |
| Primary (6) | 5(83.3%) | 1(16.7%) |  |  |
| Secondary (72) | 66(91.7%) | 6(8.3%) |  |  |
| High Grade (257) | 235(91.4%) | 22(8.6%) |  |  |
| Current Occupation (n) | Too Expensive to Take Supplements |  | p-value | r (p-value) |
|  | No n (%) | Yes n (%) |  |  |
| Housewife (84) | 77(91.7%) | 7(8.3%) | **0.1** | **0.076(0.2)** |
| Government Employed (30) | 30(100%) | 0(0.0%) |  |  |
| Private Employed (45) | 44(97.8%) | 1(2.2%) |  |  |
| Student (185) | 164(88.6%) | 21(11.4%) |  |  |
| Unemployed (4) | 4(100%) | 0(0.0%) |  |  |

### 3.10. Sociodemographic Characteristics and the Importance of Supplementation

Table 10 shows the association between sociodemographic characteristics and the perceived practical importance of taking micronutrient supplements. Overall, 95.7% of the participants found supplementation important and 4.3% did not. There was a statistically significant association between age group and perceived importance of supplementation (p = 0.001). The percentage of those who thought supplementation was important was 90.2% for women aged 16–20 years, 100.0% for women aged 21–25 years, 96.1% for women aged 26–30 years, and 100.0% for women aged >30 years. Pearson correlation also found a statistically significant positive association (r = 0.172, p = 0.001). The perceived importance of supplementation was not statistically significantly associated with marital status (p = 0.200), level of education (p = 0.500) or occupation (p = 0.200).

**Table 10:** The Association between Sociodemographic Characteristics and the Practical Importance of Taking Supplements (n=348)

| Grouped Age Years (n) | It's Important to Take Supplements |  | p-value | r (p-value) |
| --- | --- | --- | --- | --- |
|  | No n (%) | Yes n (%) |  |  |
| 16 – 20 (132) | 13(9.8%) | 119(90.2%) | **0.001** | **0.172(0.001)** |
| 21 – 25 (87) | 0(0.0%) | 87(100.0%) |  |  |
| 26 – 30 (51) | 2(3.9%) | 49(96.1%) |  |  |
| > 30 (78) | 0(0.0%) | 78(100%) |  |  |
| **Total (348)** | 15(4.3%) | 333(95.7%) |  |  |
| Marital Status (n) | It's Important to Take Supplements |  | p-value | r (p-value) |
|  | No n (%) | Yes n (%) |  |  |
| Single (209) | 13(6.2%) | 196(93.8%) | **0.2** | **0.118(0.03)** |
| Married (80) | 2(2.5%) | 78(97.5%) |  |  |
| Divorced (49) | 0(0.0%) | 49(100%) |  |  |
| Widowed (10) | 0(0.0%) | 10(100%) |  |  |
| Education Level (n) | It's Important to Take Supplements |  | p-value | r (p-value) |
|  | No n (%) | Yes n (%) |  |  |
| Illiterate (13) | 1(7.7%) | 12(92.3%) | **0.5** | **-0.026-(0.6)** |
| Primary (6) | 0(0.0%) | 6(100%) |  |  |
| Secondary (72) | 1(1.4%) | 71(98.6%) |  |  |
| High Grade (257) | 13(5.1%) | 244(94.9%) |  |  |

| Current Occupation (n) | It's Important to Take Supplements |  | p-value | r (p-value) |
| --- | --- | --- | --- | --- |
|  | No n (%) | Yes n (%) |  |  |
| Housewife (84) | 3(3.6%) | 81(96.4%) | **0.2** | **-0.082-(0.1)** |
| Government Employed (30) | 0(0.0%) | 30(100%) |  |  |
| Private Employed (45) | 0(0.0%) | 45(100%) |  |  |
| Student (185) | 12(6.5%) | 173(93.5%) |  |  |
| Unemployed (4) | 0(0.0%) | 4(100%) |  |  |

## 4. Discussion and Conclusion

### 4.1. Discussion

The aim of this study was to assess the knowledge, attitudes and practices of 348 women of reproductive age (WRA) in the Aden Governorate, Yemen toward micronutrient-rich food. Most of the participants were young and highly educated (73.9% with advanced educational levels, 53.2% students). However, economic constraints, particularly dependence on daily labor (37.9%), can greatly impact dietary choices, despite this health-literacy development profile. This finding is in line with previous evidence about the importance of education in health literacy (21).

Overall, participants had a relatively good knowledge of micronutrient rich foods and 77.9% knew that micronutrients are important for health. Specific food sources were well recognized, particularly liver/red meat (92.8%), eggs (91.4%), fruits (90.5%) and dairy products (89.4%). However, only 29.6% recognized iodized salt as a relevant source of micronutrients, indicating an important knowledge gap. This pattern is consistent with evidence that more complex nutritional concepts are less well known than practical dietary sources (24). The low recognition of iodized salt is a public health concern because of its role in preventing iodine deficiency especially in women and children (25,26).

Knowledge about the effects of micronutrient deficiencies was less complete: only 26.2% were able to identify all of the major health problems related to deficiencies, while 6.9% were unable to identify any. Similar constraints in micronutrient knowledge and practices have been reported among adolescent girls in India, highlighting the need for targeted nutrition education (27). Interestingly, high knowledge of pregnancy-related nutritional needs (96.5%) and dietary diversity (95.2%) did not translate into consistently optimal dietary practices and supports the knowledge-practice gap observed in the present study (12).

Economic and social factors seemed to be major barriers. Over two-thirds of the participants (70.6%) considered healthy food to be too expensive, consistent with evidence from Yemen that found affordability to be a major determinant of food consumption (28). Similarly, 32.7% indicated social influence in cooking, supporting the evidence that women’s dietary practices are shaped by social, economic and knowledge related factors (29). However, the 59.5% preference for traditional foods could be an opportunity for culturally appropriate interventions promoting nutrient-rich local foods.

Another gap in perception was seen on the purpose of micronutrient-rich foods. Almost half the participants (46.0%) reported these foods as important for infant health, while only 18.1% reported them as important for their own health. This highlights the necessity of addressing women’s nutritional well-being other than their reproductive function, especially in light of the maternal health challenges in Yemen (30).

Nutritional vulnerability was also indicated by dietary practices. While 76.2% reported eating three meals a day, daily intake of eggs (36.2%), meat/poultry/fish (32.2%) and especially fruits (13.2%) were low. These results are in line with previous evidence of inadequate daily consumption of different food groups in women (24). Regular micronutrient supplementation was also infrequent, with only 9.8% reporting regular use. This is concerning given the nutritional burden in Yemen and national commitments to provide iron and folic acid supplementation (31).

The most reported barrier to nutritious food access was high food cost (43.4%), which confirms the importance of addressing economic constraints as well as nutrition education (29,30). Only 3.7% of participants identified poor food-preparation knowledge as a barrier, but this may not fully reflect the knowledge–practice gap, especially since nutrition education should include food preparation, storage, hygiene and meal planning (19).

Sociodemographic analysis revealed significant association of knowledge with education, age, marital status and occupation. Education was most strongly positively correlated with knowledge (r = 0.514, p < 0.001), consistent with prior evidence (32,33). Participants who were younger and students also had higher knowledge. There was a weak positive correlation between age and knowledge (r = 0.162, p = 0.002). This confirms findings on the advantages of nutrition education in younger populations (27).

Only 8.3% considered supplements to be expensive while 95.7% agreed on their importance. Supplement affordability varied significantly by age but not by education or occupation. Comparable evidence suggests that supplement cost may be a relatively minor barrier in some populations (35). The high perceived importance of supplementation given the variable knowledge further indicates a knowledge–attitude gap that may be addressed by targeted education and behavior-change interventions (38,33).

### 4.2. Conclusion

Women of reproductive age in Aden showed relatively good knowledge of micronutrients and generally positive attitudes toward supplementation; however, poor dietary practices and affordability are important challenges. Therefore, interventions should include complementary nutrition education, access to affordable and micronutrient-rich foods, and appropriate supplementation.

### 4.3. Limitations

This is a cross-sectional study and causality cannot be determined. The lack of biochemical measures limited the objective assessment of micronutrient status. Further, data collection at a particular point in time may not have sufficiently considered seasonal variations in dietary practices.

## Declarations

### Ethical considerations and consent to participate

Participants were recruited from 01/06/2026 to 01/09/2026. Before participating all participants gave verbal informed consent. The participants were informed about the aim and procedures of the study and participation was voluntary. Confidentiality and privacy were maintained during the study. Ethical approval for the study was obtained from the University Research Committee with approval number (UST-ETH-2026-0015). Informed consent was obtained from all participants before data collection. Participants were voluntary. The participants were informed about the purpose and procedures of the study and during the process of research confidentiality and privacy were maintained. The data were collected and used only for research purposes.

### Consent for publication

This manuscript does not present any individual participant identifying information or identifiable personal data.

### Data and material availability

The datasets generated and/or analyzed during the current study are available from the corresponding author on reasonable request.

### Authors Contributions

Mohammed A. A. Al-Baghdadi: Conceptualization, Supervision, Methodology, Review. Shifaa Suhail Mohammed: Collected data, investigated, analyzed and drafted the original manuscript. Shrook Jalal Ahmed Data collection, investigation, analysis and original drafting. Mohammed Ali Khalifa: Conceptualization, methodology, statistical analysis, interpretation, original draft writing and review/editing. All authors approved the final version of the manuscript.

## Acknowledgments

The authors would like to thank the women who participated in this study for their valuable time and contribution. We also appreciate the support and cooperation of the staff of the participating primary health care and maternal and child health centers in Aden Governorate, Yemen, in the data collection. We are grateful to all the people who contributed to the successful completion of this study.

## Appendix 1: Ethical Approval Letter

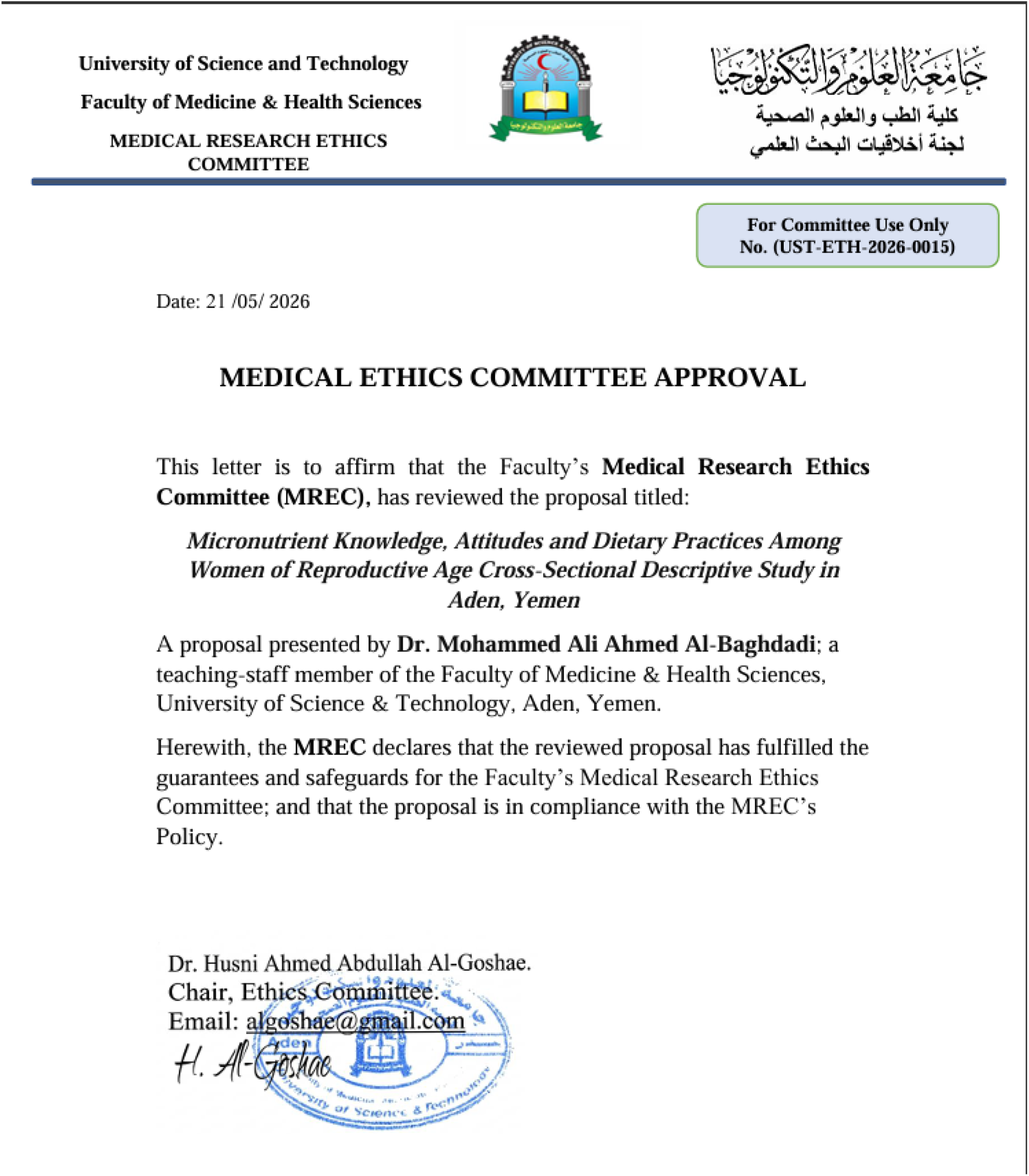

## Appendix 2: Data Collection Form

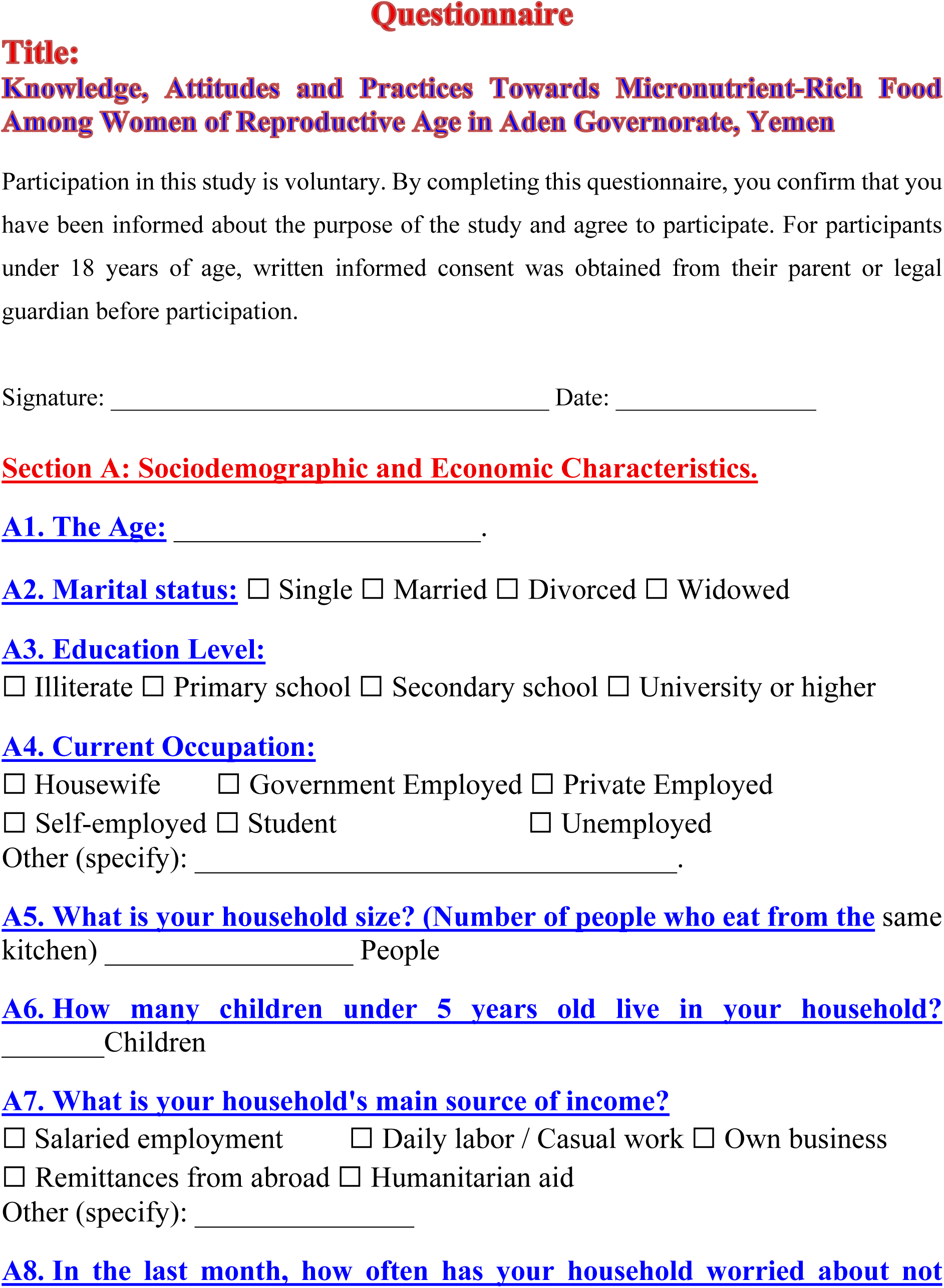
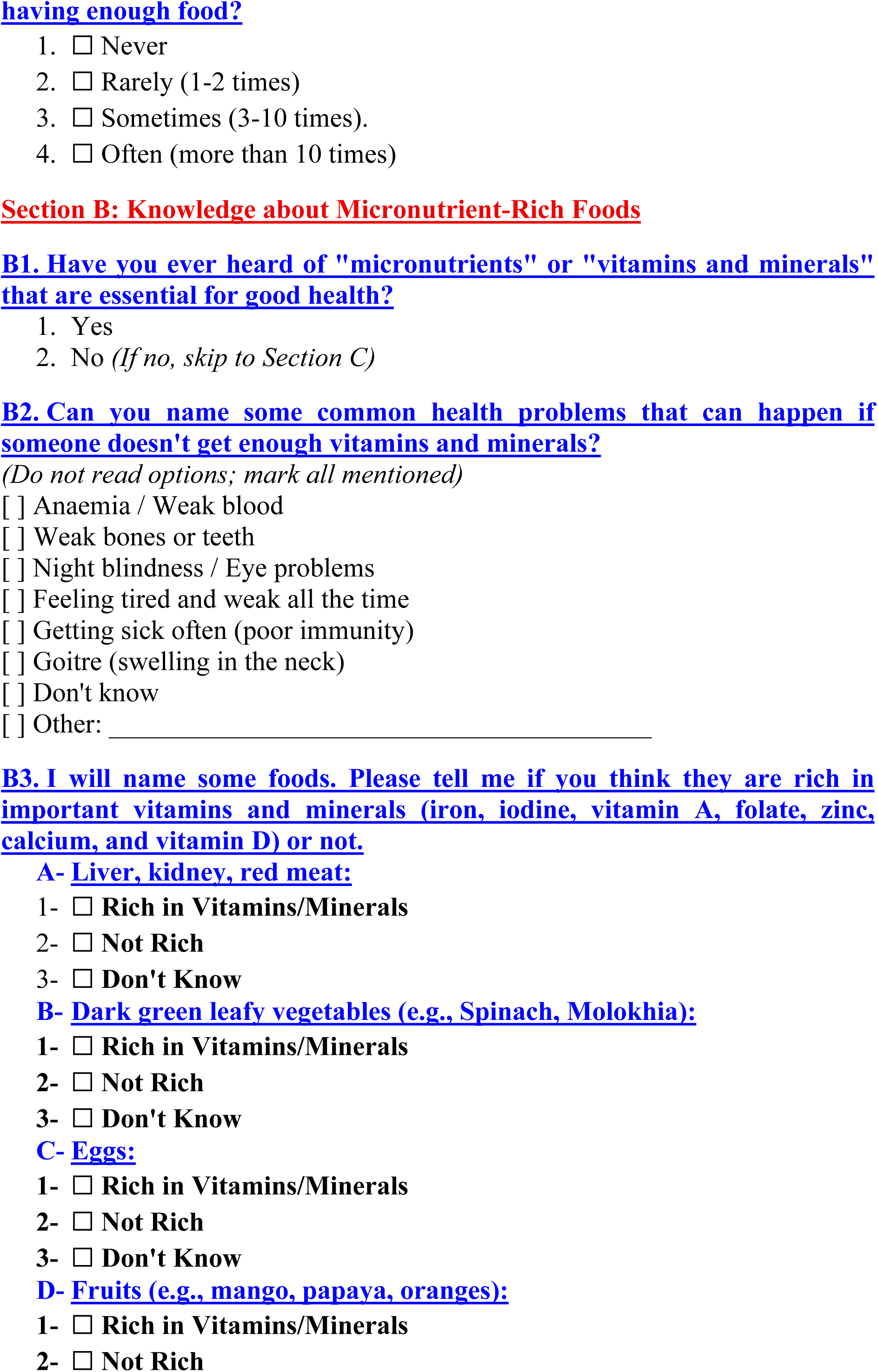
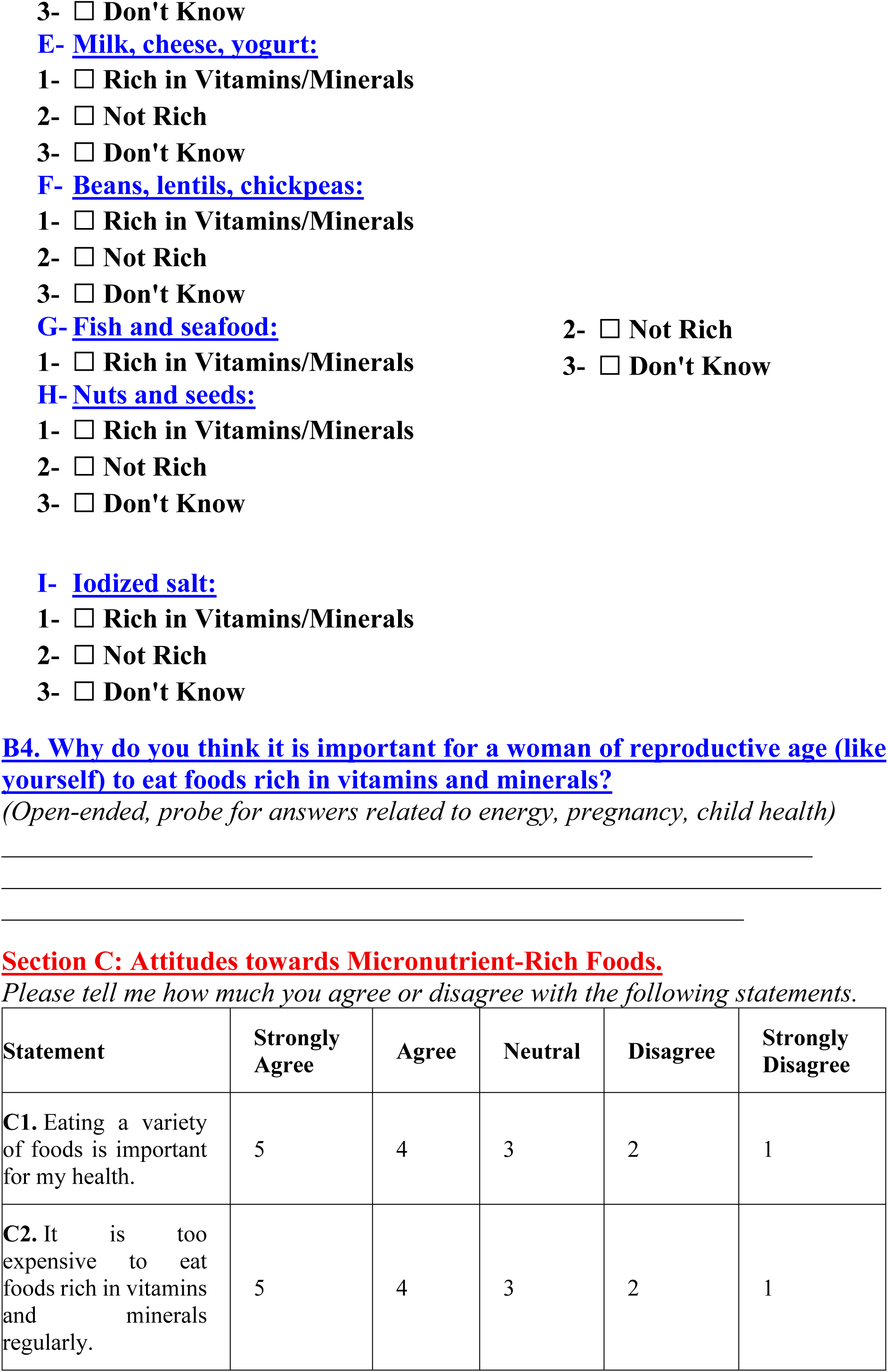
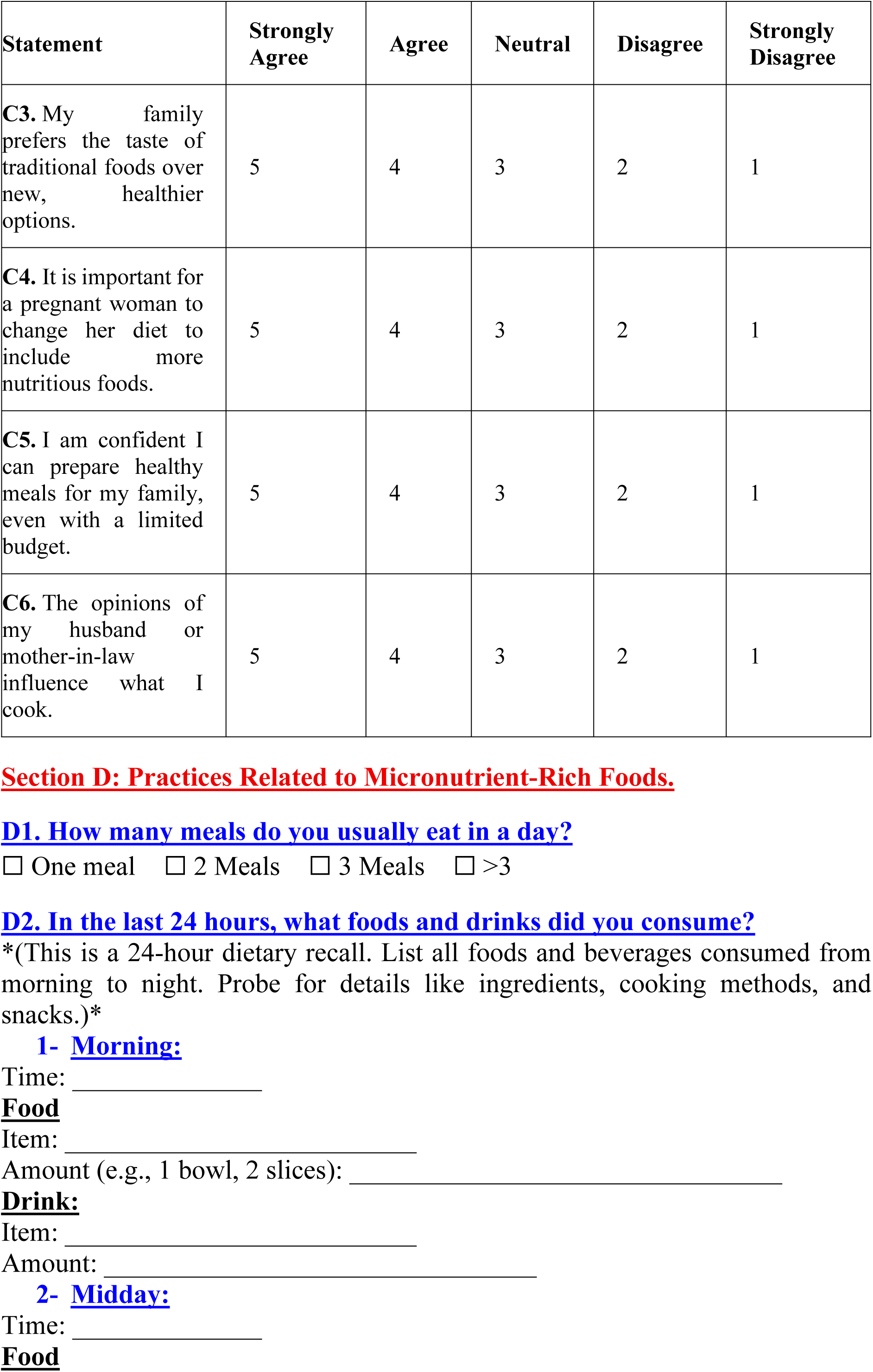
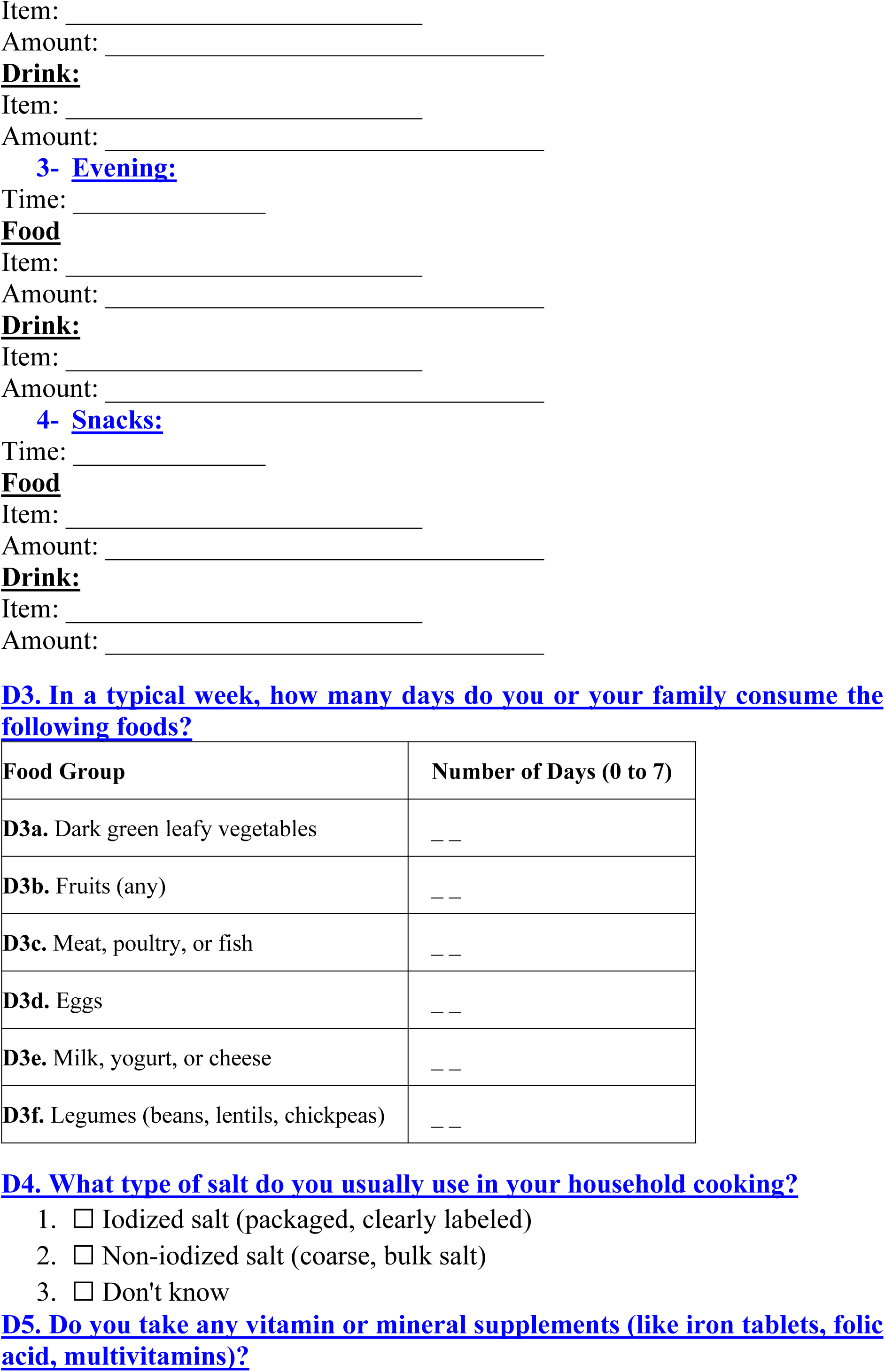
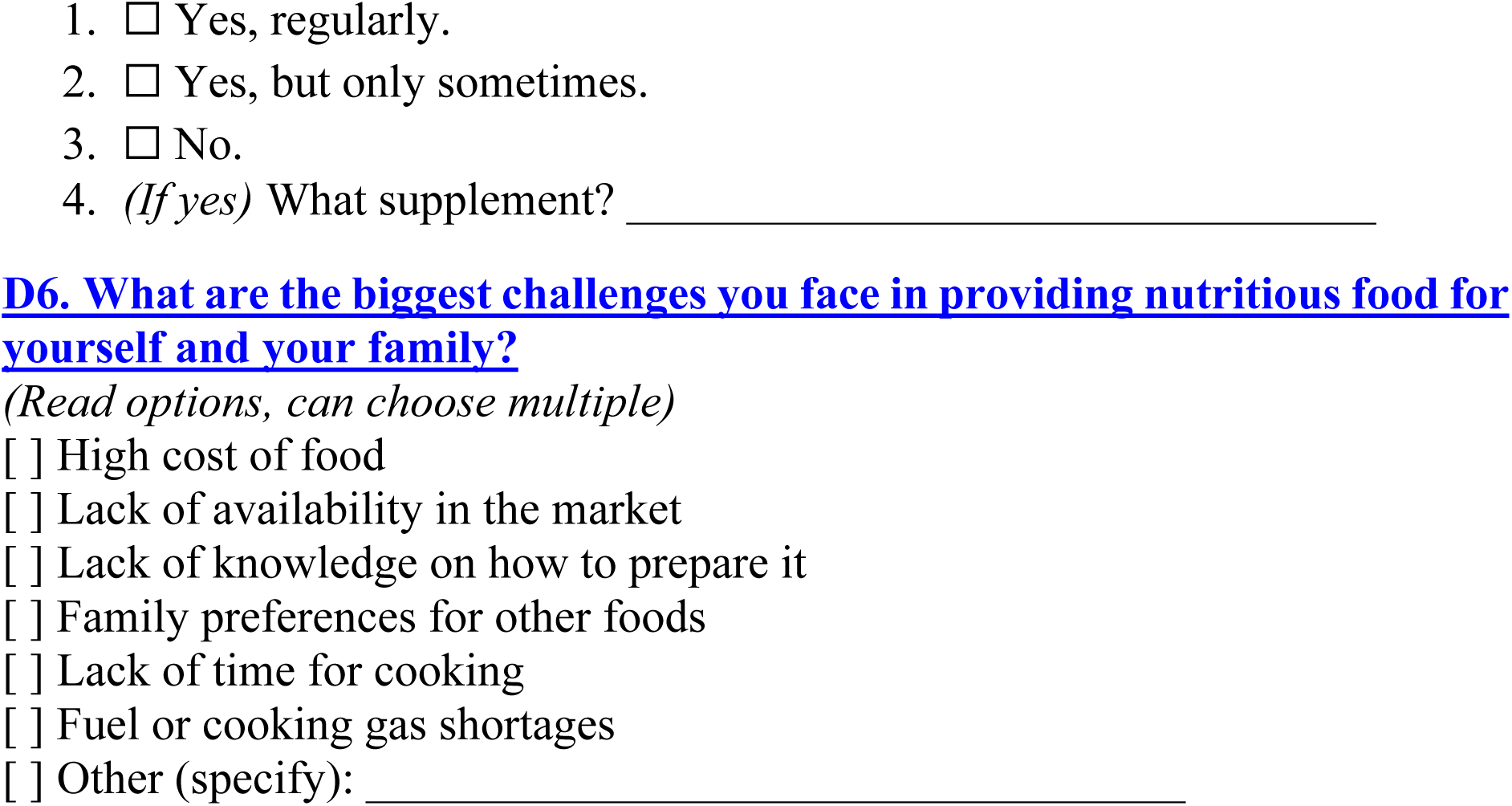

